# Longitudinal Cardiorespiratory Wearable Sleep Staging in the Home

**DOI:** 10.1101/2025.08.27.25334365

**Authors:** Shaun Davidson, Jonathan Carter, Emily C Stanyer, Rachel Sharman, Cristian Roman, Simon D Kyle, Lionel Tarassenko

## Abstract

There is a growing interest in performing automated, longitudinal tracking of sleep in the home environment using wearables and machine learning. Wearables such as smart watches or chest patches can be comfortably worn for long periods of time, and cardiorespiratory waveforms measured by these wearables combined with machine learning models to estimate sleep state. However, these machine learning models are typically trained and tested on retrospective data from sleep labs, where cardiorespiratory waveforms are monitored using bulky, specialised equipment rather than wearables. As such, these models have yet to be validated in their intended scenario of use: longitudinal, wearable sleep monitoring in the home.

This paper establishes and validates a pipeline for longitudinal cardiorespiratory sleep monitoring in the home using data from the RESTORE study. In RESTORE, 17 participants with a sleep-related condition (insomnia and depressive symptoms) underwent a sleep-related clinical intervention (sleep restriction therapy). Participants simultaneously wore a low-density home electroencephalogram device, allowing for expert, manual sleep staging using brain activity, as well as a wearable chest patch, allowing for wearable monitoring of cardiorespiratory waveforms. Both devices were worn by participants for 10 nights while undergoing treatment at home. A state-of-the-art cardiorespiratory sleep staging model, combining transformer and convolutional neural networks, was then tuned and tested on the wearable data using leave-one-subject-out-cross-validation.

After transfer learning, the cardiorespiratory sleep staging model had an accuracy of 77.1% and Cohen’s Kappa of 0.679 for four-class sleep staging using the wearable data from the RESTORE study. Further, the model was able to accurately track sleep and sleep-derived metrics longitudinally while participants underwent sleep restriction therapy. These results represent one of the first direct demonstrations of the potential for wearable, cardiorespiratory sleep staging to track longitudinal, clinically relevant changes in sleep in individuals undergoing a sleep-related intervention in the home.

## 1. Introduction

There is a growing body of literature on the association between sleep and health. Chronic sleep deprivation is known to result in neural, molecular, and immune changes that contribute to cardiovascular disease development [1]. Further, the timing and relative amount of particular sleep states is known to be associated with susceptibility to, and progression of, a diverse range of conditions including dementia, diabetes, and depression [2, 3, 4]. The continuous monitoring of sleep patterns has the potential to provide early indication of disease progression and treatment response, as well as potential targets for intervention [4, 5].

Gold-standard sleep monitoring is performed using polysomnography (PSG), which typically includes monitoring an individual’s brain activity via electroencephalography (EEG), eye movement via electrooculography (EOG), muscle movement via electromyography (EMG), heart activity via electrocardiography (ECG), and breathing via Impedance Pneumography (IP) or nasal airflow in a dedicated sleep lab. Each 30 seconds of sleep, typically referred to as an ‘epoch’, is then manually labelled by an expert scorer as wake, Rapid Eye Movement (REM), Non-REM 1 (N1), Non-REM 2 (N2), or Non-REM 3 (N3 or slow wave) sleep [6]. PSG is expensive and demanding of expert time for manual labelling of sleep stages [7]. PSG also conventionally requires a subject to sleep in an unfamiliar lab environment while wearing obtrusive monitoring equipment, potentially disrupting a participant’s sleep (sometimes referred to as the ‘first night effect’) [8]. As such, while sleep research using PSG remains the gold standard and has produced compelling findings in small, targeted cohorts [4, 5], it is not a feasible approach for large-scale, longitudinal monitoring of sleep.

Recent advances in wearable technology and machine learning have the potential to address this issue [9]. There are now a variety of wearable devices that can provide a subset of PSG or PSG-adjacent waveforms in a continuous and minimally disruptive fashion. These include wearable EEG-based devices such as the Dreem headband, which exhibit good performance but have trade-offs in terms of price, tolerability, and ease-of-use [10]. A less disruptive approach to wearable sleep monitoring is to analyse cardiorespiratory waveforms acquired with wrist- or chest-worn wearables, which are typically unobtrusive and can be worn continuously for multiple days [11]. While manual sleep staging is performed primarily using the EEG, EOG, and EMG, cardiorespiratory waveforms are associated with sleep behaviour via the autonomic nervous system, and machine learning models can be trained to leverage this association for automated sleep staging [12, 13]. Although photoplethysmography (PPG) measured at the wrist is a sensing technology used extensively in commercial devices, an advantage of chest-worn wearables is that they can provide a direct measurement of not only cardiac but also respiratory signals, a combination that has been shown to improve sleep staging performance [14].

A variety of machine learning models for cardiorespiratory sleep staging have been reported in the literature [12, 13, 15]. These models are typically trained and evaluated on large, open access, retrospective PSG datasets such as those provided in the National Sleep Research Resource (NSRR) [16]. Given the relatively low incidence and low inter-rater agreement associated with N1 sleep, it is common for these cardiorespiratory sleep staging models to combine N1 and N2 sleep into a single ‘light sleep’ class, enabling them to perform four-class sleep staging (i.e, wake, N1/N2, N3, REM) [14, 15]. Recent advances incorporating the transformer architecture have demonstrated near expert-level sleep scoring using ECG and IP waveforms from sleep lab PSG datasets (Cohen’s Kappa of 0.78 and accuracy of 84.8% in [14]). However, these methods have yet to be directly validated in their intended scenario of use - longitudinal, chest-worn wearable data in the home environment. This environment presents several potential challenges, including differences in signal quality between wearable and PSG-derived ECG, as well as the fact that most chest wearables do not directly provide an IP waveform.

Highlighting this challenge, a recent publication [17] investigated wearable sleep staging using the Physionet DREAMT dataset [18]. This dataset contains expert scored PSG data alongside PPG data from a research grade wearable smart watch (Empatica E4) for 100 participants (1 night per participant). [18] report a Cohen’s Kappa of 0.553 after fine tuning/transfer learning for three-class sleep staging (i.e., wake, Non-REM, and REM, with no differentiation between N1/N2 and N3 sleep), outperforming previous state-of-the art models including SleepPPGNet [15] when fine tuned and tested on the same dataset. It is notable here that the Cohen’s Kappa reported for SleepPPGNet after fine tuning on the DREAMT dataset (0.426) is significantly lower than that reported for retrospective PSG test data (0.74 in [15]), underlining the challenges of translating such models from sleep lab PSG waveforms (in this case, PPG from a clinical grade enclosed finger probe, as in the NSRR datasets [16]) into the home environment (in this case, PPG from a research grade wearable smart watch in DREAMT).

In this paper, we establish a pipeline for extracting a respiratory signal from a wearable chest patch accelerometer and then performing cardiorespiratory sleep staging using a state-of-the-art transformer model. We assess the performance of this pipeline against manual, expert sleep labelling using a low-density EEG/EOG/EMG device in the home environment. Our study cohort includes longitudinal monitoring of participants with a sleep-related condition (insomnia disorder and depressive symptoms [5]) undergoing a sleep-related intervention (sleep restriction therapy [19, 20]). As such, this study allows for the assessment of the model’s ability to track sleep in a scenario that closely resembles a potential use case for a wearable cardiorespiratory sleep staging system.

## 2. Materials and Methods

### 2.1. The RESTORE Study

Wearable cardiorespiratory data and expert sleep labels were drawn from the “examining the mechanisms of sleep RESTriction therapy fOR insomnia in people with dEpressive symptoms” (RESTORE) study [21]. The primary outcome of the RESTORE study was the feasibility of serial home EEG monitoring in participants undergoing Sleep Restriction Therapy (SRT), as assessed by the proportion of EEG recordings that were scorable and yielded usable data, as well as the feasibility of the study for participants, and acceptability of the EEG equipment both qualitatively and quantitatively. A secondary aim was to investigate the physiological mechanisms underlying SRT’s potential to ameliorate depressive symptoms [19, 20]. The RESTORE study was conducted in Oxfordshire, UK and was approved by the Medical Sciences Interdivisional Research Ethics Committee (MS IDREC) at the University of Oxford (Approval reference: R91701/RE001). Informed consent was obtained from all participants prior to their involvement. Full details of the study protocol are available at [21].

Participants were recruited through community advertising and assessed for eligibility via an online questionnaire and interview [21]. Overall, 17 participants with insomnia disorder (Sleep Condition Indicator (SCI) *<*= 16), depressive symptoms (Hospital Anxiety and Depression Scale (HADS-D) *>*= 8), and who were not taking medication that could affect their sleep, were recruited. Once recruited, participants underwent a 6-week study protocol (shown in Fig. 1), which included a 2-week baseline period and 4-week SRT period. Participants recorded daily sleep diaries and underwent daily affect assessments (e.g., Positive and Negative Affect Schedule (PANAS)) throughout the study period.

**Figure 1.**
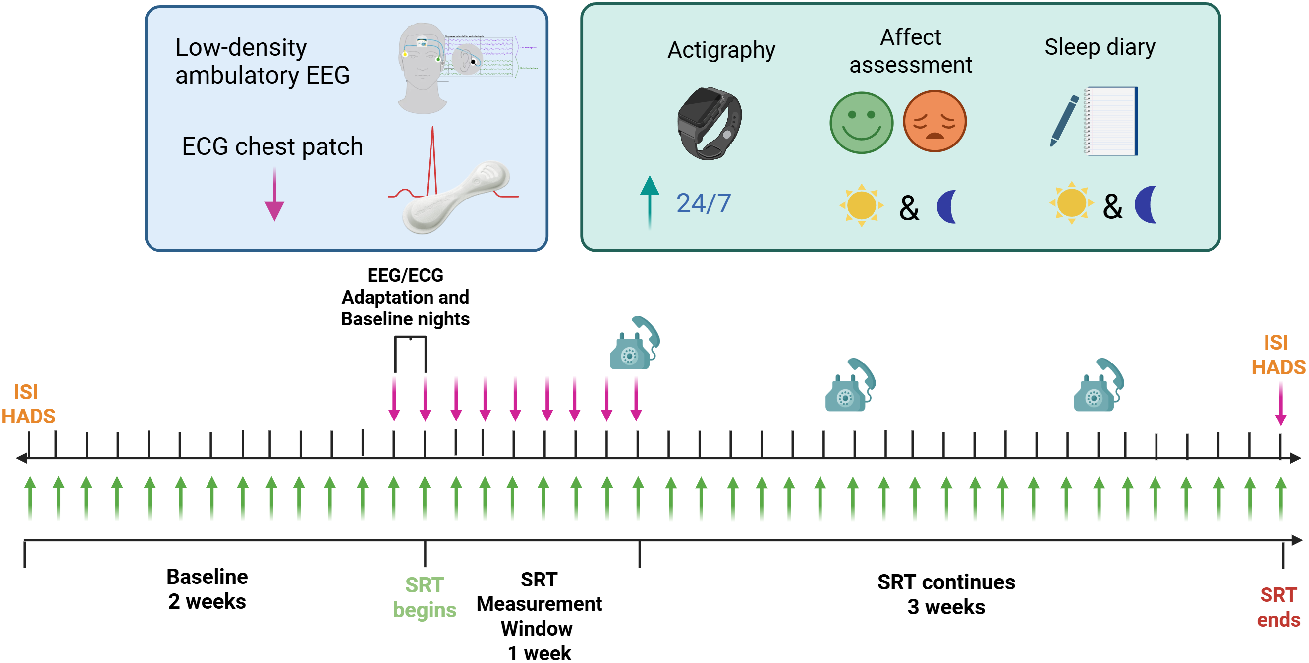
Summary of the RESTORE study protocol. ISI is the Insomnia Severity Index, and HADS the Hospital Anxiety and Depression Scale.

Each participant also underwent 10 nights of EEG sleep monitoring (1 adaptation night, 1 baseline night directly prior to beginning SRT, the 7 initial nights of SRT, and 1 night at the end of the SRT protocol). During these nights, participants wore a low-density EEG device (HomeSleepTest (HST) device, Somnomedics, similar to [22, 23]), which included a forehead (FPz) EEG electrode (with the output signal filtered between 0.3 and 35 Hz, according to AASM standards [24]), left and right EOG electrodes, and EMG/reference (M1/mastoid) electrode, each sampled at 256 Hz. Simultaneously, participants also wore an adhesive 7-day chest-patch (VitalPatch, VitalConnect), part of an in-house wearable remote monitoring system [25], which directly records continuous ECG and three-axis accelerometer measurements at 125 Hz. The chest patch was initially applied to the participant by the research team prior to the baseline recording session, with subsequent reapplications of the chest patch after 7 days of monitoring performed at home by the participant. The HST device was self-applied by the participant prior to each night of EEG sleep monitoring. All monitoring occurred in the participant’s home environment.

At the conclusion of the RESTORE study, sleep stages for all non-adaptation nights (i.e., up to 9 nights per participant) were labelled by a blinded expert scorer (ECS) using data from the HST and DOMINO v3.0.0.8. Scoring followed the AASM guidelines version 3 [24] with consideration to the montage (e.g., single forehead FPz EEG and mastoid EMG).

### 2.2. Extracting a Respiratory Signal

While the chest patch directly records ECG sampled at 125 Hz and reports an estimated respiratory rate every 4 seconds, it does not output a respiratory waveform. Instead, we extract a respiratory waveform (ACC-RESP) from the three-axis accelerometer signal (which is sampled at 125 Hz) provided by the chest patch. To achieve this, we use a modified version of the recursive method described in [26]. With this method, a respiratory signal is extracted by projecting the accelerometer signal onto the plane perpendicular to gravity and then using Principal Component Analysis (PCA) to identify the major axis of chest respiratory movement. This process effectively extracts a respiratory waveform, but emphasises different components of respiratory chest wall movement (i.e., those perpendicular to gravity) depending on whether the individual being monitored is lying supine or in a lateral position. In contrast, a thoracic IP waveform, such as that typically available in PSG studies, employs an impedance band which detects overall chest wall movement, regardless of body position. To extract a respiratory waveform from the accelerometer data that more closely resembles a thoracic IP waveform and captures respiratory chest wall movement, regardless of whether an individual is lying supine, prone, or in a lateral position, we developed a modified approach which avoids assuming that the direction of chest wall respiratory movement is perpendicular to gravity. This approach was implemented as described in the supplementary material.

### 2.3. wav2sleep Model

For determining sleep states from the continuous ECG and ACC-RESP signals, we use the state-of-the-art deep learning architecture developed in [14], which combines transformer and Convolutional Neural Network (CNN) components. This architecture has been shown to outperform existing cardiorespiratory sleep staging methods across a variety of signal input modalities, achieving a Cohen’s Kappa of 0.78 and accuracy of 84.8% when using ECG and respiratory (thoracic IP) input waveforms. wav2sleep is designed to simultaneously classify sleep state for periods of up to 10 hours (i.e, 1200 epochs), and consists of three components:

- **Signal Encoders**: These consist of a series of residual blocks [27] which extract features from the raw input signals (ECG and IP) using a configuration based on SleepPPGNet [15]. Each block consists of three convolutional layers followed by a pooling operation, as shown in Fig. 2. Separate encoders are used for each signal modality, facilitating different input sample rates (parameters shown in Table 1). Once features are extracted, they are grouped per epoch by performing a reshape operation, and then fused into a reduced set of 128 features per epoch using a time-distributed dense operation (parameters in Table 1).
- **Epoch Mixer**: This consists of two transformer encoder blocks [28] that fuse the features extracted by the signal encoders for each signal modality into a single representation. This encoder attends over the entire night’s (i.e., 1200 epochs) worth of extracted features, allowing for the modelling of long-range feature interactions and dependencies across the entire night of sleep using positional encoding. A simplified representation of an epoch mixer encoder block is shown in Fig. 2 and supplemental parameters are provided in Table 1.
- **Sequence Mixer**: This is a dilated CNN that takes the unified representation output by the epoch mixer and generates sleep state labels for each epoch. A sequence mixer block is shown in Fig. 2 and parameters are provided in Table 1. Dilated convolutions increase the receptive field of a CNN, allowing for long-range feature interactions and influences to be modelled. These long-range interactions are important to consider when labelling sleep state, where long-range factors such as time since the beginning of sleep or position within a sleep cycle have known influences on sleep behaviour.

**Table 1.** wav2sleep model parameters and layer input/output dimensions from [14]. Block architectures are shown in Fig. 2.

| Model Component | Parameters | Output Dim |
| --- | --- | --- |
| Raw Input, ECG | 34.13 Hz | 1228800, 1 |
| Raw Input, IP | 8.53 Hz | 307200, 1 |
| Signal Encoder, ECG | i in [16, 16, 32, 32, 64, 64, 128, 128] | 4800, 256 |
| Signal Encoder, IP | i in [16, 32, 64, 64, 128, 128] | 4800, 256 |
| Reshape (ECG) | Group features per epoch | 1200, 1024 |
| Reshape (IP) | Group features per epoch | 1200, 1024 |
| Time-Distributed Dense (ECG) | Dense (1024 in, 128 out) | 1200, 128 |
| Time-Distributed Dense (IP) | Dense (1024 in, 128 out) | 1200, 128 |
| Epoch Mixer (ECG+IP) | 2 Encoder Blocks, Dropout 0.1 | 1200, 128 |
| Sequence Mixer | i in [1, 2, 4, 8, 16, 32] × 2 dilation | 1200, 128 |
| Sleep State Prediction | Dense (128 in, 4 out) | 1200, 4 |

**Figure 2.**
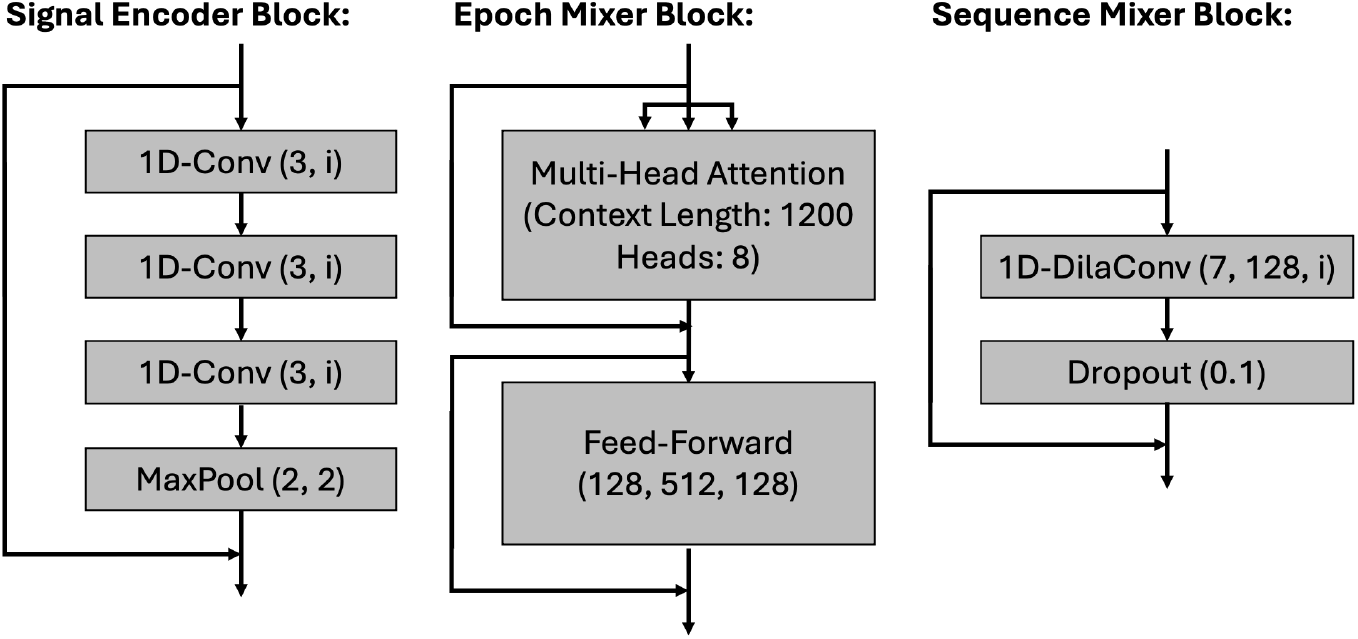
Example signal encoder, epoch mixer, and sequence mixer blocks. For the signal encoder, each 1D-Conv layer has a kernel size of 3 and number of features (‘i’) specified in Table 1. For the sequence mixer, each 1D-Dila(ted) Conv layer has a kernel size of 7, 128 features, and dilation factor specified in Table 1.

### 2.4. Transfer Learning

Due to the domain shift between PSG and wearable ECG, and the ACC-RESP signal extracted from the chest patch accelerometer not directly corresponding to the thoracic IP signal in the labelled PSG data used to train wav2sleep, we perform transfer learning to enable the model to adapt to the changed signal modalities. As a starting point, we use the pre-trained wav2sleep model checkpoint provided by [14], which was trained on over 10,000 expert-labelled overnight PSG recordings across 7 of the constituent databases in the NSRR [16]. Each participant in RESTORE was included in the analysis if good quality HST (i.e., containing any data considered scoreable by expert annotators) and chest patch data (i.e., no overnight gaps in data greater than 30 minutes in duration) were available for at least 75% of their study nights.

Given the relatively small number of participants in the RESTORE study, we implement Leave-One-Subject-Out-Cross-Validation (LOSOCV) when assessing model performance. For each participant in RESTORE, this entails making a given participant the test subject, splitting data from the other participants into training and validation cohorts, performing transfer learning, and then evaluating model performance on the test individual’s data. In line with [14], the model was trained using the AdamW optimiser and Cross Entropy loss with a batch size of 16. Transfer learning was performed using a fixed learning rate of 10^-5^, with training stopped if the validation loss did not decrease for 5 consecutive epochs. The model weights were then reverted to those from the model state with the lowest validation loss for evaluation. Overall performance metrics are reported for the accuracy of sleep staging across all test subjects, with each participant serving as the test subject in turn.

## 3. Results

### 3.1. Study Cohort

Overall, 17 participants were recruited for the RESTORE study, the demographics of whom are shown in Table 2. Participants were mostly female (76.5%) and of white British ethnicity (70.6%). Table 2 also summarises cohort sleep metrics computed at baseline from the HST device, as well as baseline depression (HADS) and insomnia (ISI) scores.

**Table 2.** Demographics of the RESTORE cohort.

| **Demographic** | **Value** |
| --- | --- |
| Age (years), mean (SD) | 38.2 (13.9) |
| Age Range | 19 - 62 |
| Female, n (%) | 13 (76.5) |
| **Ethnicity, n (%)** |  |
| White, British | 12 (70.6) |
| White, Other | 1 (5.9) |
| Asian, Any | 0 (0.0) |
| Black, Any | 1 (5.9) |
| Mixed | 2 (11.8) |
| Other | 1 (5.9) |
| **Baseline Sleep, mean (SD)** |  |
| Total Sleep Time (mins) | 414.9 (70.9) |
| Time in Bed (mins) | 461.2 (72.6) |
| Sleep Efficiency (%) | 89.8 (4.5) |
| Sleep Onset Latency (mins) | 16.7 (14.1) |
| **Insomnia & Depression, mean (SD)** |  |
| ISI | 16.8 (3.7) |
| HADS | 10.0 (2.8) |

Of the 17 participants recruited for the study, 13 had good quality HST and chest patch data available for at least 75% of their study nights, giving a total of 105 nights of data for analysis (with each participant contributing 7 - 9 nights of data). As shown in Table 3, this corresponds to 84,209 epochs sleep data labelled by a single expert sleep scorer, with an average of 6,478 labelled epochs per participant. Note that any epochs manually labelled as ‘artefact’ by the expert scorer were excluded when computing performance metrics. To perform LOSOCV over each of the 13 participants, data from the remaining 12 participants was randomly split into training and validation sets, as described in Section 2.4. In each case, data from 10 participants was used for training and data from the other 2 participants for validation.

**Table 3.**
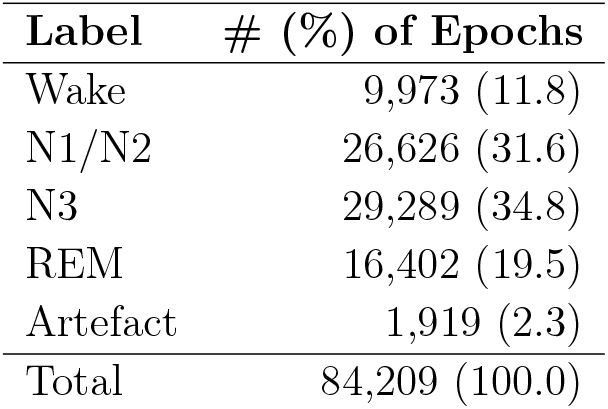
Distribution of Expert-Labelled Epochs in the RESTORE Dataset.

| **Label** | **# (%) of Epochs** |
| --- | --- |
| Wake | 9,973 (11.8) |
| N1/N2 | 26,626 (31.6) |
| N3 | 29,289 (34.8) |
| REM | 16,402 (19.5) |
| Artefact | 1,919 (2.3) |
| Total | 84,209 (100.0) |

### 3.2. wav2sleep Model Performance - Without Transfer Learning

Fig. 4 shows confusion matrices summarising the LOSOCV performance of the wav2sleep model for cardiorespiratory sleep staging without transfer learning on the 105 nights of data from 13 RESTORE participants included in this analysis. It is notable here that, even without transfer learning, both models show reasonably good agreement with manual expert sleep scoring (accuracy 68.0% and Cohen’s Kappa 0.554 for the model using ECG+ACC-RESP as inputs, and accuracy 66.9% and Cohen’s Kappa 0.542 for the model using only ECG as an input). This is despite considerable domain shift between the PSG data used to train the model and the wearable sensor data used in this study. Further, performance is still improved by adding our accelerometer-derived respiratory waveform despite substantial differences in data acquisition and pre-processing compared to a thoracic IP signal (as employed in the training data for the wav2sleep model [14]). Across both models, agreement between expert scoring and model-based cardiorespiratory sleep staging is generally good for wake (82.32% - 85.45%), N1/N2 sleep (81.21% - 86.61%), and REM sleep (77.25% - 78.45%), but less so for N3 sleep (40.25% - 41.75%).

**Figure 4.**
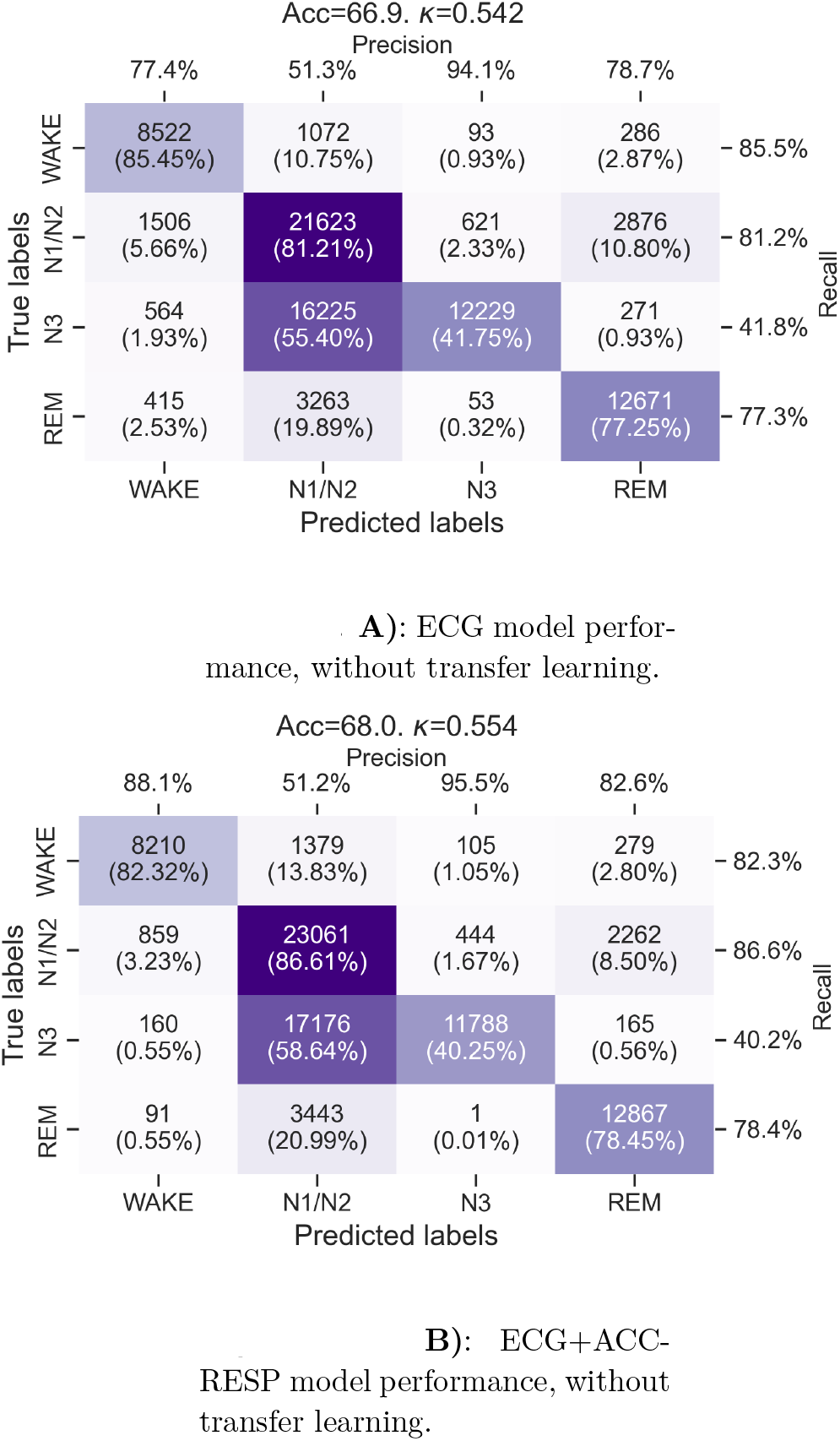
Confusion matrices for the sleep staging performance of the wav2sleep transformer model without transfer learning. **(A)** Using only ECG from the chest patch as an input. **(B)** Using ECG and accelerometer-derived respiratory waveform from the chest patch as inputs.

### 3.3. wav2sleep Model Performance - With Transfer Learning

Fig. 5 shows confusion matrices summarising the performance of the wav2sleep model for cardiorespiratory sleep staging after transfer learning is performed. Overall model performance is significantly improved compared compared to before transfer learning (Accuracy from 68.0% to 77.1%, Cohen’s Kappa from 0.554 to 0.679 for ECG+ACC-RESP, and Accuracy from 66.9% to 75.3%, Cohen’s Kappa from 0.542 to 0.655 for ECG only). Once again, the model incorporating ECG+ACC-RESP as inputs outperforms the model using only ECG as an input. For both models, accuracy across classes is considerably more consistent compared to before transfer learning, ranging from 74.09% - 83.00% for the ECG+ACC-RESP model and 71.20% - 80.42% for the ECG only model.

**Figure 5.**
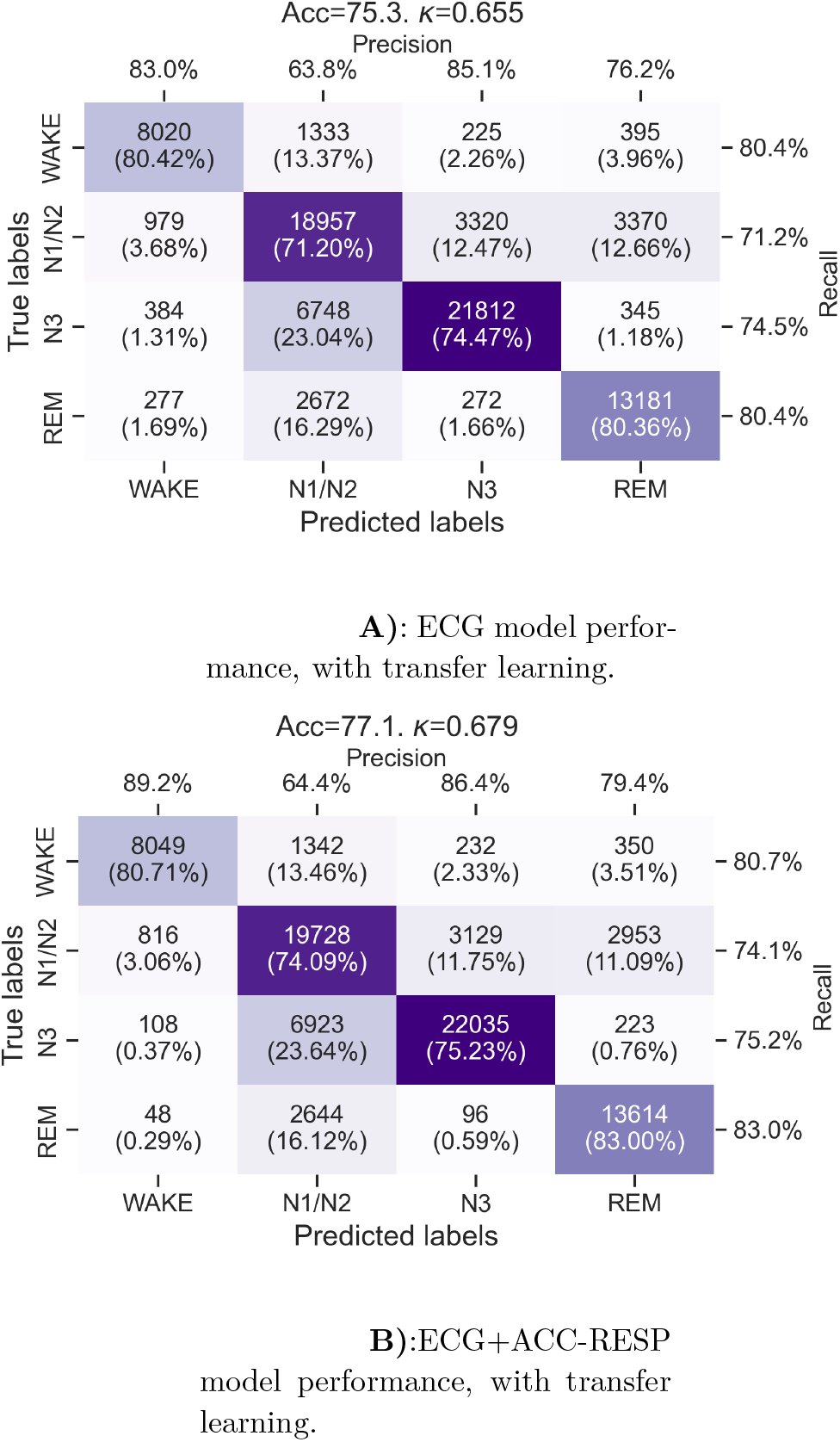
Confusion matrices for the sleep staging performance of the wav2sleep transformer model with transfer learning. **(A)** Using only ECG from the chest patch as an input. **(B)** Using ECG and accelerometer-derived respiratory waveform from the chest patch as inputs.

Fig. 6 shows an example comparison of model-derived hypnograms before and after transfer learning with manual expert EEG sleep staging. In Fig. 6 it can be observed that the major shift the transfer learning process introduces is to increase the model detected incidence of N3 sleep (as compared to N1/N2 sleep) to better align with manual expert EEG labels.

**Figure 6.**
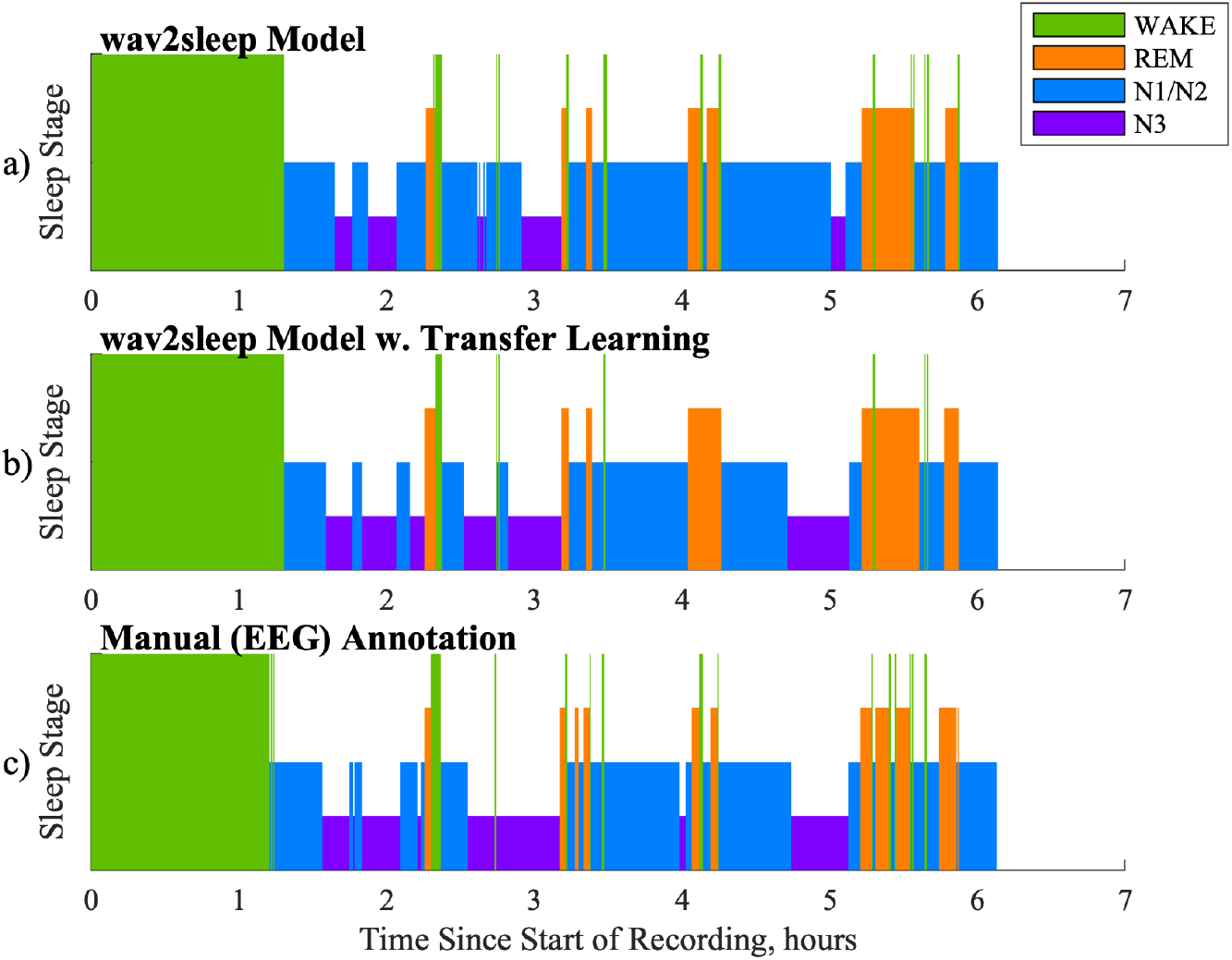
Example comparison of hypnograms between: a) wav2sleep model before transfer learning; b) wav2sleep model after transfer learning; c) Manual Expert (EEG) annotation of sleep.

Fig. 7 shows an example of longitudinal trends in total sleep time, %N1/N2, %N3, and %REM sleep for a participant with good quality data for all 9 recorded nights during the RESTORE protocol. In Fig. 7 it can be observed that model trends in total sleep time align well with expert labelling regardless of transfer learning, but that the transfer learning process notably improves the ability of the model to accurately track longitudinal trends in the relative incidence of different sleep states while participants are undergoing the RESTORE protocol. Fig. 8 highlights that transfer learning improves the ability of the model to longitudinally track changes in N1/N2 sleep and N3 sleep across the entire RESTORE cohort.

**Figure 7.**
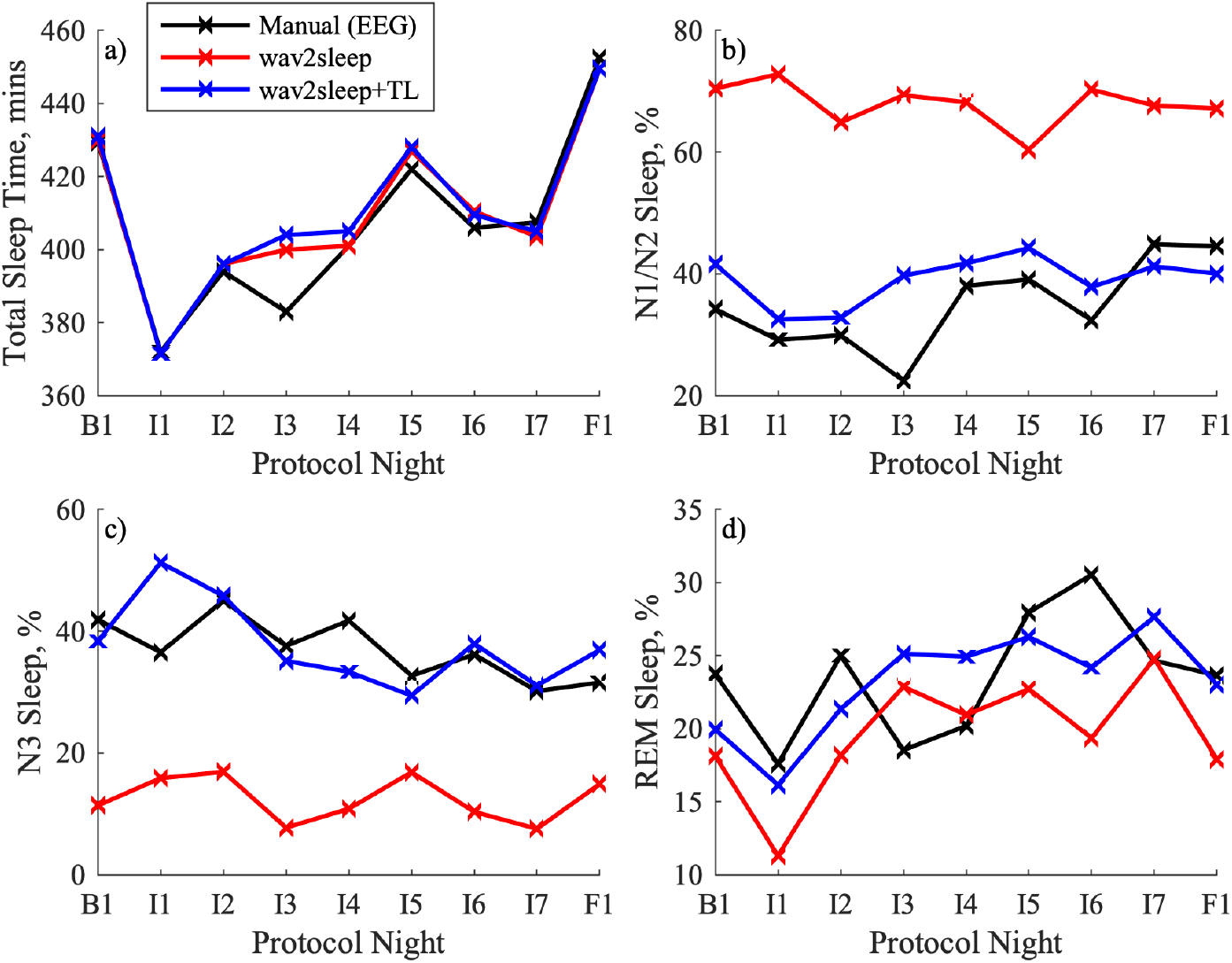
Example comparison of longitudinal trends for a RESTORE participant in: (a) Total Sleep Time; b) %N1/N2 Sleep; c) %N3 Sleep; d) %REM Sleep. B1 denotes baseline, I1-I7 denote intervention, and F1 denotes follow up nights.

**Figure 8.**
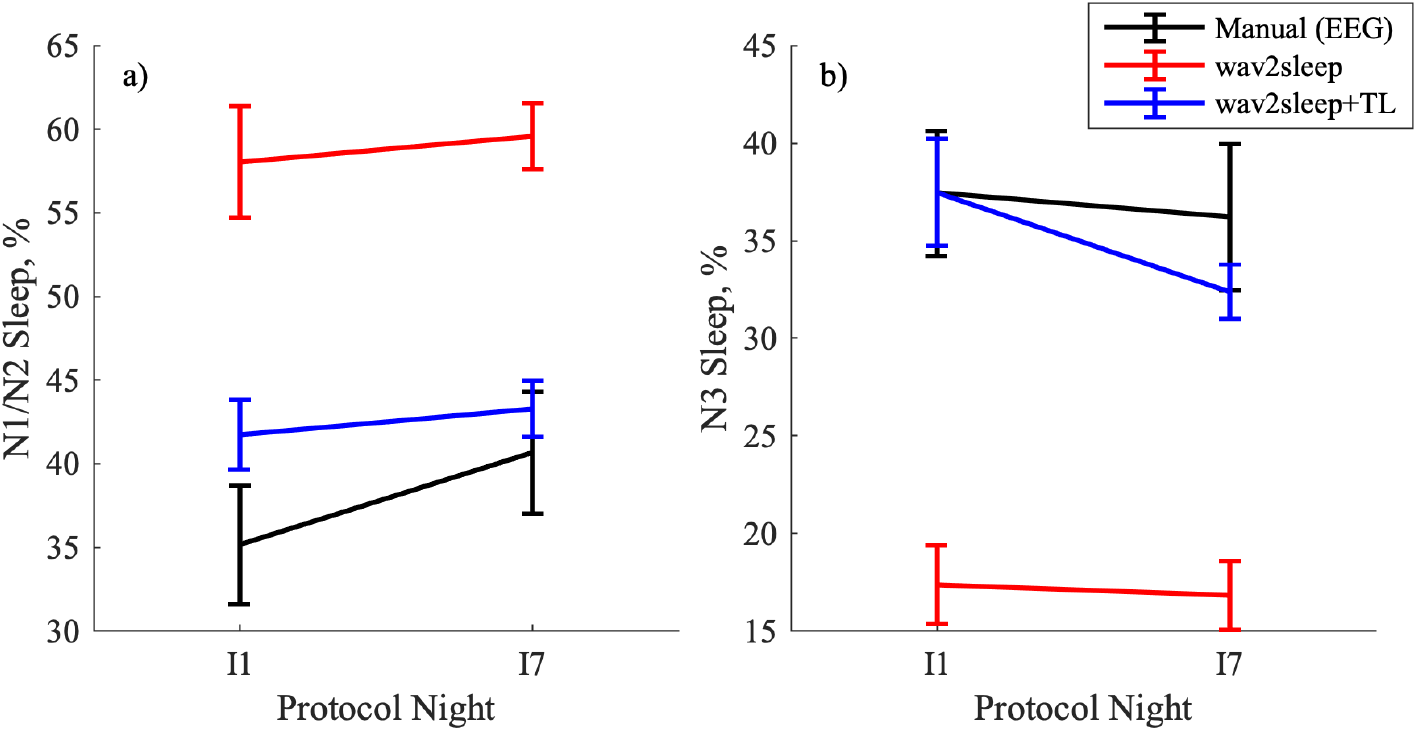
Comparison of sleep data from two nights across the RESTORE cohort for: a) %N1/N2 Sleep; b) %N3 Sleep. I1 and I7 denote the first and last nights of intervention, respectively. Error bars show standard error of the mean.

## 4. Discussion

Prior to transfer learning, the transformer model exhibits reasonable agreement with manual, expert sleep staging using a low-density HST device, with an accuracy 66.9% and Cohen’s Kappa of 0.542 (using only ECG as an input, Fig. 4) or an accuracy of 68.0% and Cohen’s Kappa of 0.554 (using ECG and ACC-RESP as inputs, Fig. 4). The model exhibits very good performance for classifying wake, N1/N2, and REM sleep (accuracies of 77.25% - 86.61%), but notably worse performance for classifying N3 sleep (accuracies of 40.25% - 41.75%), with N3 sleep primarily misclassified as N1/N2 sleep (55.40% - 58.64%). Both overall and per-class sleep staging accuracies are lower than those reported for the retrospective PSG test set in [14], with a slight reduction in wake accuracy (90.78% to 83.32% - 85.45%) and N1/N2 accuracy (88.96% to 81.21% 86.61%), a moderate reduction in REM accuracy (89.53% to 77.25% - 78.45%), and a notable reduction N3 accuracy (72.24% to 40.25% - 41.75%). The slight reduction in accuracy for wake and N1/N2 sleep is likely attributable to differences in signal quality between PSG and wearable cardiorespiratory sensors, as well as the shift from a respiratory waveform derived from a thoracic impedance band to one derived from a 3-axis accelerometer, without any transfer learning to account for this change in modality.

While these factors are also likely to contribute to the greater reductions in accuracy observed for REM and N3 sleep, there are other factors to consider regarding these two sleep states. With regards to the moderate decrease in REM accuracy compared to that reported in [14], it is well established that ocular artefacts can contaminate frontal EEG signals, and that this effect can be particularly significant for the FPz forehead electrode due to its proximity to the eyes [29]. With regards to the notable drop in N3 accuracy compared to that reported in [14], it is important to note that the NSRR databases used to train and test the transformer model label N3 sleep using the C4/A1 or C3/A2 central electrodes (the ‘acceptable’ AASM electrode configuration), as opposed to the frontal F4/A1 or F3/A2 electrodes (the ‘ideal’ AASM electrode configuration) [16, 24]. It is additionally important to note that slow waves are known to exhibit greater amplitude in frontal EEG leads due to their association with the frontal regions of the brain [30]. Given that the FPz forehead electrode montage utilised by the HST device is even further forward than the ‘ideal’ F4/A1 or F3/A2 electrodes, it is unsurprising that the application of the AASM 75 µV threshold (which has well-established limitations [31]) by expert scorers to identify slow waves from this electrode montage leads to a relatively increased scoring of N3 sleep, especially when compared to a model trained using sleep stage labels from central electrodes. Notably, the incidence of EEG-labelled N3 sleep reported in this study cohort is also significantly greater than that reported in prior meta-analyses for individuals with insomnia and depression [32].

Transfer learning can be used to address differences both in input signal modality (i.e., the shift in respiratory signal from thoracic IP to ACC-RESP) or class labelling (i.e., the shift from N3 sleep labelled using the 75 µV threshold on a central (C4/A1) electrode to N3 sleep identified using a higher-amplitude forehead (FPz) electrode montage). Figs. 5 and 5 show the impact of transfer learning for models using ECG only and ECG+ACC-RESP and as inputs, respectively. Overall model performance is significantly improved, with accuracy increasing from 68.0% to 77.1% and Cohen’s Kappa from 0.554 to 0.679 for the model using ECG+ACC-RESP as inputs, and accuracy increasing from 66.9% to 75.3% and Cohen’s Kappa from 0.542 to 0.655 for the model using only ECG as an input. After transfer learning, the additional accuracy gained by including ACC-RESP as a model input increases from 1.1% to 1.8%, potentially reflecting the model adapting to the ACC-RESP waveform, having been trained on the thoracic IP waveform present in the NSRR datasets. Accuracy across sleep stages is considerably more consistent after transfer learning, ranging from 74.09% (for N1/N2 sleep) to 83.00% (for REM sleep) for the ECG+ACC-RESP model. Model accuracies for N3 and REM sleep increase at the expense of a slight decrease in accuracy for wake and a moderate decrease in accuracy for N1/N2 sleep. In particular, the accuracy for scoring N3 sleep increases from 40.25% to 75.23%, with N3 misclassification as N1/N2 decreasing from 58.64% to 23.64%.

When compared to the results in the retrospective PSG dataset in [14], model accuracies for wake and REM sleep are similar, while model accuracy for N1/N2 sleep is notably lower (88.96% in [14] compared to 74.09% in Fig. 5) and N3 sleep accuracy slightly greater (72.24% in [14] compared to 75.23% in Fig. 5). The likely explanation for this shift is the relatively greater incidence of N3 sleep in the RESTORE dataset (Table 3) due to scoring N3 sleep, with the accompanying trade-off in N1/N2 sleep scoring, using the frontal FPz electrode montage. This, in turn, results in N3 sleep having a relatively greater contribution to the cross-entropy loss used to train the model, resulting in the model placing an increased emphasis on identifying N3 sleep at the expense of N1/N2 sleep after transfer learning.

The example hypnograms in Fig. 6 support these points, showing the model’s ability to classify wake and REM accurately even prior to transfer learning (importantly capturing the short initial REM cycle at 2.25 hours - the timing of which is relevant for the monitoring of conditions such as depression [5], as present in the RESTORE cohort). The effect of transfer learning here is the increase in the relative incidence of model labelled N3 sleep as compared to N1/N2 sleep, better aligning hypnograms with expert sleep scoring using the HST device’s FPz electrode.

The example longitudinal trends for a RESTORE participant in Fig. 7 highlight the future potential of this work. In Fig. 7, B1 reflects the baseline night, I1-I7 the intervention nights (i.e., Sleep Restriction Therapy (SRT)), and F1 the follow-up night. The wav2sleep model tracks total sleep time equally well before and after transfer learning, capturing the decrease in total sleep time when beginning SRT, followed up a gradual increase as the participant’s sleep schedule adjusts during the first week of SRT, and finally an increase in total sleep time during follow-up after 4-weeks of SRT. After transfer learning, the model is similarly able to capture trends in N1/N2 sleep (slight initial decrease followed by gradual increase during intervention), N3 sleep (gradual decrease), and REM sleep (initial decrease, increase during intervention, decrease during follow-up). These results are complemented by those in Fig. 8, which show that transfer learning notably improves the ability of the model to track changes in N1/N2 and N3 sleep during SRT across the RESTORE cohort. Importantly, these results represent one of the first direct demonstrations of the potential for wearable, cardiorespiratory sleep staging to track longitudinal, clinically relevant changes in sleep in individuals undergoing a sleep intervention in the home, rather than in a sleep lab.

This study is, to our knowledge, the first to investigate longitudinal, wearable cardiorespiratory sleep staging in the home environment. This investigation is further strengthened by a comparison with manual, expert EEG sleep scoring while participants with a sleep-related condition (insomnia and depressive symptoms) undergo an intervention directly targeting sleep (sleep restriction therapy). As previously discussed, existing literature on wearable cardiorespiratory sleep staging typically uses waveforms from retrospective, open access PSG datasets as a surrogate for wearable data [13, 33]. While this provides a valuable resource for initial model training and benchmarking, such PSG-derived cardiorespiratory data are not fully representative of wearable cardiorespiratory data gathered in the home, as highlighted by the reduction in performance of state-of-the-art models such as SleepPPGNet reported in [17] when transitioned from PSG to wearable datasets.

A further important takeaway from the results in [17] is the advantages of performing sleep staging using chest-based (as compared to wrist-based) wearable waveforms. PPG from wrist-worn wearables is a popular signal modality for sleep staging in commercial devices due to its low burden. However, the Cohen’s Kappa of 0.553 for three-class sleep staging using smart watch PPG reported in [17], outperforming existing state-of-the-art methods using the same input signal, is notably less than the Cohen’s Kappa of 0.679 for four-class sleep staging reported here using the ECG and an accelerometer-derived respiratory waveform in a cohort of individuals undergoing a sleep intervention. Importantly, chest worn wearable waveforms contain significantly more respiratory information than wrist-worn PPG, both indirectly through respiratory modulation of the ECG and directly through the accelerometer-derived respiratory waveform. Four-class sleep staging has significant clinical advantages over three-class sleep staging as it allows the differentiation of N3 sleep, which has a variety of important clinical associations [2], from N1/N2 sleep.

### 4.1. Limitations

An important limitation of this study is the limited number of participants (13) and diversity (mostly white, female participants) in the RESTORE cohort. This limitation should be, however, be considered in the context of the longitudinal nature of the RESTORE study, which encompassed a total of 105 nights (82,290 epochs) of manually scored sleep epochs (i.e., similar in size to the 100 nights of data in the Physionet DREAMT dataset [18]). Nevertheless, a future study incorporating a larger, more diverse cohort of participants would provide broader validation for the methodology developed in this manuscript.

A further limitation was the use of a limited montage, low-density EEG device (Somno-HST), as opposed to 19-lead PSG recommended by AASM scoring guidelines, for manual EEG sleep staging. The use of this low-density device was important in facilitating the study being performed in a longitudinal fashion on consecutive nights in the home environment, which would be difficult to accomplish using gold standard PSG due to the high cost and disruption associated with such an approach [7]. It is worth noting that the low-density device does provide AASM standard left and right EOG, as well as single-lead EEG from the FPz electrode and EMG from the M1 electrode, and that sleep staging was performed manually by an expert sleep scorer. Further, a similar EEG/EOG/EMG device from Somnomedics was previously validated against gold standard PSG in [22], and general performance of home sleep monitoring devices using the FPz electrode in [23]. Regardless, direct validation of the cardiorespiratory sleep staging pipeline in a non-longitudinal study against gold-standard PSG would certainly complement the results presented in this paper.

With regards to the RESTORE study methodology, it is worth noting that no exclusion criteria were set for participants taking medication which might affect their cardiovascular system. Such medication could potentially affect the coupling between the autonomic nervous system and sleep states that cardiorespiratory sleep staging relies upon. Further, it is worth noting that participants self-applied the HST device and chest patch (after being initially shown how to apply this), which could lead to variable signal quality depending on placement. This self application does, however, mean that the dataset more closely resembles a true ‘use case’ scenario for this technology.

### 4.2. Conclusion

Overall, this paper develops a pipeline for performing wearable cardiorespiratory sleep staging in the home environment. This pipeline involves deriving a respiratory waveform from a chest worn accelerometer (as a surrogate for thoracic IP acquired from a chest band) and combining it with wearable ECG to perform cardiorespiratory sleep staging using a state-of-the-art deep learning model integrating transformer and CNN components. This pipeline is validated in a cohort of participants with a sleep related condition (insomnia and depressive symptoms) undergoing a sleep-related clinical intervention (sleep restriction therapy) in the home environment. This work thus represents one of the first validations, and an important step towards the realisation, of this promising new technology in its intended use case - wearable, longitudinal sleep monitoring in the home environment.

## Data Availability

The raw data supporting the conclusions of this article will be made available by the authors, without undue reservation.

