## Supplementary material for "Longitudinal Cardiorespiratory Wearable Sleep Staging in the Home": Supplemental Material.pdf

### Extracting a Respiratory Signal from the Accelerometer

The ACC-RESP signal was extracted using the following process:

(i) Low-pass filter the three-axis accelerometer signal using a 3rd order Butterworth filter with a cut-off frequency of 3.5 Hz.

(ii) Linearly re-sample the filtered accelerometer signal down to 16 Hz, giving the three-axis accelerometer signal $\vec{a}$.

(iii) Compute the gravity vector at the sample point $n$ as the recursive, rolling average of the three-axis accelerometer signal using equation 1.

$$\vec{g}[n] = \alpha_h \vec{a}[n-1] + (1-\alpha_h)\vec{g}[n-1] \tag{1}$$

Where $\vec{g}$ is the gravity vector (which is initialised with the value [0 0 1]) and $\alpha_h$ is a coefficient described in Table 1. The gravity vector is used to determine whether movement is occurring using equation 2.

$$m[n] = \|\vec{a}[n] - \vec{g}[n]\| > 0.06\|\vec{g}[n]\| \tag{2}$$

Where $m[n]$ is a movement flag, which is applied for the subsequent 2.5 seconds (15 samples) when movement occurs and affects the coefficients described in Table 1.

(iv) For each of the three accelerometer axes, remove the DC component, thereby extracting the respiratory component of the signal, by subtracting the 5-second (80-sample) moving mean, giving the three-axis respiratory signal $\vec{h}$. This is in contrast to the method described in [1], in which the gravity vector is used to determine the direction of respiratory motion, giving a two-axis signal.

(v) Recursively normalise the respiratory signal $\vec{h}$ using equation 3.

$$\vec{u}[n] = \frac{\vec{h}[n]}{p[n]} \tag{3}$$

where $\vec{u}$ is the normalised respiratory signal and $p$ is a rolling, recursive norm computed using equation 4.

$$p[n] = \alpha_n \|\vec{h}[n]\| + (1 - \alpha_n)p[n-1] \tag{4}$$

where $\alpha_n$ is a coefficient described in Table 1 and $p$ is initialised by taking the mean norm of the first 10 samples of $\vec{h}$

(vi) Low-pass filter the normalised respiratory signal $\vec{u}$ using a 3rd order Butterworth filter with a cut-off frequency of 0.8 Hz, giving the output signal $\vec{v}$.

(vii) Perform recursive PCA on the three-axis respiratory signal $\vec{v}$ (as opposed to the two-axis signal in [1]) using equation 5.

$$\hat{r}[n] = \vec{w}[n] \cdot \vec{v}[n] \tag{5}$$

where $\hat{r}[n]$ is the accelerometer-derived respiratory signal (ACC-RESP), and the direction of principal component $\vec{w}$ is approximated recursively using equation 6.

$$\vec{w}[n+1] = \eta\hat{r}[n]\vec{v}[n] + (1 - \eta\hat{r}[n]^2)\vec{w}[n] \tag{6}$$

where $\eta$ is a learning rate described in Table 1, and $\vec{w}$ is arbitrarily initialised as [1 0 0].

(viii) Adjust the sign of the ACC-RESP signal to ensure that inspiration and expiration have consistent directions (positive and negative, respectively). This is achieved using the skewness $s$, which is recursively approximated using equation 7.

$$s[n] = \alpha_s\hat{r}[n]^3 + (1 - \alpha_s)s[n-1] \tag{7}$$

where $\alpha_s$ is a coefficient described in Table 1. If the skewness falls below a threshold of 5e-3 at a sample point $n$, the direction of inspiration and expiration are assumed to be incorrect, and the skewness $s[n]$ and projection axis $\vec{w}[n]$ at that sample point are inverted.

An example segment of night time chest-patch accelerometer data with the associated accelerometer-derived respiratory waveform is shown in fig. 1.

**Table 1.** Movement-affected coefficients for extracting a respiratory waveform from three-axis accelerometer, as established in [1]. The greater 'Body Movement' values allow the recursive method to adapt more rapidly after a potential change in orientation by a participant.

| Parameter | Normal Value | Body Movement Value |
|---|---|---|
| $\alpha_h$ | 0.03 | 0.30 |
| $\alpha_n$ | 0.01 | 0.10 |
| $\alpha_s$ | 7e-5 | 7e-4 |
| $\eta$ | 8e-4 | 8e-3 |

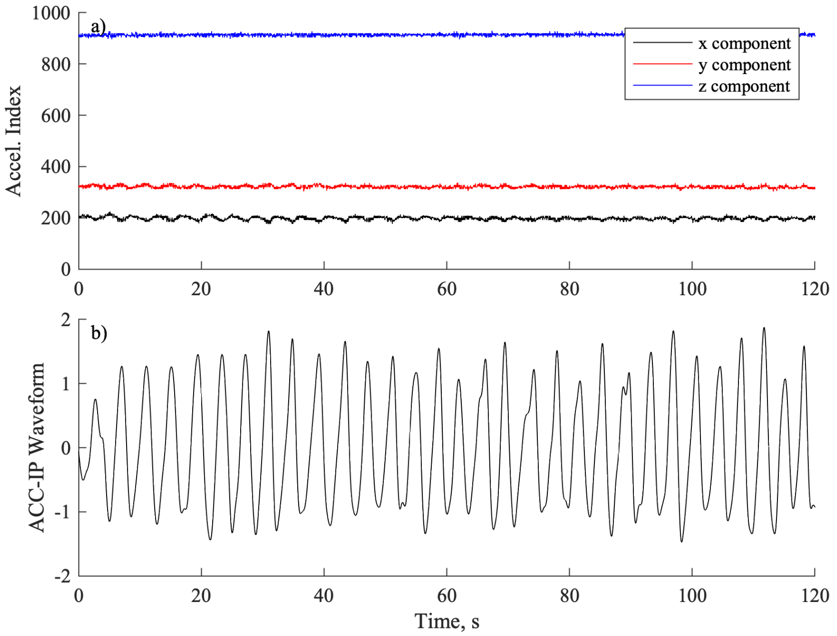


**Figure 1.** Example 120 second segment of overnight waveforms for: a) chest-patch derived 3-axis accelerometry at 125 Hz; b) accelerometer-derived respiratory waveform (ACC-RESP).
